# The relationship between ethnicity and stone composition in a large multi-ethnic London NHS Trust

**DOI:** 10.1101/2021.11.24.21266803

**Authors:** S Vaggers, R Warner, L Forster, Z Ali, P Pal, S Graham

**Affiliations:** Whipps Cross University Hospital; Royal London Hospital

## Abstract

**Purpose:** Few studies have examined kidney stone composition of an ethnically diverse group living in the same location, we aimed to study this in an ethnically diverse NHS trust.

**Methods:** We reviewed all patients (n=312) with laboratory stone analysis and compared their ethnicity with their stone composition.

**Results:** Using a Chi Squared analysis there was no significant difference between stone composition in different ethnic groups (p=0.07). Uric acid stones were more common in the White-other group at 22.0% compared to 10.3% for White British, 5.2% for Asian and 9.52% for Black patients. Calcium oxalate stone were more common in the Asian population with 71.9% and Black population at 76.1%, compared with 56.7% in the White British population and 52.6% in the White-other population. Calcium phosphate stones were found commonly in the White British population (26.8%) compared to 18.6% for White-other, 16.7% for Asian and 9.5% for Black patients. Cystine and Struvite stones were found at low levels of between 0-3.4% in each group. Repeat stone formers with calcium oxalate, uric acid or cystine stones formed the same stone again 100.0% of the time. The odds for the Black population having a stone analysed (OR 0.62, CI 0.39-0.97, p=0.04) was significantly lower than the local population, and for the Asian population this was significantly higher (OR 1.31, CI1.05-1.62, p=0.01),

**Conclusion:** Uric acid stones are found more frequently in the White-other population and calcium oxalate stones are found more frequently in the Asian and black population. However, these results were not statistically significant. The odds ratio of having a stone was significantly higher in the Asian population and lower in the Black population.

## Introduction

There is a wide variation in the prevalence of different stone types across the world^2, 3-6^. For example, Calcium oxalate stones are the most common, but their prevalence varies, being significantly higher in the Asian subcontinent (75-93%[1–5]) compared to the Western world (65.98- 67%^19-20^). Uric acid stones are found at relatively high proportions specific populations 15.8-50%^3,4^ and far lower 8-10.8% across most other studies[6, 7]. There may be a wide range of diets, fluid intake, medical conditions, medications, climate and genetics which are influencing the composition of these stones.

It is not known whether it is patient’s ethnicities or environmental factors specific to their location which have a greater influence on kidney stone composition. There are a very limited number of studies comparing stone composition of an ethnically diverse group living in the same location[8, 9]; most of these have been done in the United States.

The area of East London covered by this NHS trust has a particularly wide variation in ethnicities.

We were interested to discover whether patients with different ethnicities would retain a variation in the frequency of different stone types similar to Western Europe or similar to their specific ethnicity. With this information we may be able to identify whether environment or genetics are more important in determining the chemical composition of a patient’s kidney stone.

## Methods

### Study design

We performed a retrospective cohort study of all patients with laboratory stone analysis in the largest NHS trust in the UK in 2017, Barts Health NHS Trust. This NHS trust provides care for an estimated 2.6 million people living in east London[10]. The policy is to send stone analysis on every patient where stone retrieval is achieved.

Ethical approval was not required in accordance with the NHS research and ethics committee. The study involved analysis of existing data with no additional data collected with no allocation to an intervention and no randomisation. No patients were excluded from the study.

### Patient demographics and stone characteristics

Patient demographics (including ethnicity) and kidney stone composition were determined from hospital electronic records. Ethnicity was self-determined by the patient according to national codes recommended by the Office for National Statistics[11]. Stone size obtained from radiology reports on the picture archiving and communication system (PACS). Stone composition was designated as the chemical compound with the highest percentage.

### Previous stone analysis

We reviewed each patient’s electronic records for the past 10 years. For every patient with a previous laboratory stone analysis their previous stone composition was noted.

### Statistics

The data was analysed using Microsoft Excel using a binomial test for the frequency of stones in male and female patients and the Chi squared test was used to effect of ethnicity on stone type. Patients whose ethnicity was unknown and stones whose composition were unknown were still included in separate categories. Graphs were creating in excel. Odds ratios were used to determine the likelihood of different groups being found in our stone population compared to the local population. A p value of less than 0.05 was set as statistically significant.

## Results

### Demographics

312 patients had a stone sample collected and laboratory stone analysis out of a total of 630 patients undergoing percutaneous nephrolithotomy, ureteroscopy or extracorporeal shockwave lithotripsy. 156 (31.1%) of these patients were White British, 59 (18.9%) were White – any other White background, 96 (30.8%) Asian, 21 (6.7%) Black, and 39 (12.5%) other - any other ethnic group/ unspecified/ refused (Table 1). The median age of the patients was 48 with an interquartile range of 37-60. The male:female ratio was 2.39:1.

**Table 1.** Descriptive statistics of the ethnicity of our population

| Ethnicity | Number of patients | Percentage of patient of this ethnicity in our study |
| --- | --- | --- |
| **White** |  |  |
| White British | 156 | 31.1% |
| White other | 59 | 18.9% |
| **Mixed/Multiple ethnic groups** |  |  |
| Any other Mixed/Multiple ethnic background: | 3 | 1.0% |
| **Asian/ Asian British:** |  |  |
| Indian | 16 | 5.1% |
| Pakistani | 19 | 6.1% |
| Bangladeshi | 38 | 12.2% |
| Chinese | 3 | 1.0% |
| Any other Asian background | 20 | 6.4% |
| **Black/African/Caribbean/Black British** |  |  |
| African | 8 | 2.6% |
| Caribbean | 8 | 2.6% |
| Any other Black/African/Caribbean background | 5 | 1.6% |

| Other ethnic group |  |  |
| --- | --- | --- |
| Any other ethnic group | 12 | 3.8% |
| Other: not stated | 18 | 5.8% |
| Refused | 6 | 1.9% |

### Stone characteristics

The average size of the stone was 9.92mm (SD 7.12).

Calcium oxalate was the commonest stone type in all groups: (range 52.5-76.9%, overall: 64.4%) with calcium phosphate being the second most common (range 9.5-26.8%, overall: 20.2%). Uric acid stones were seen in 9.6% of patients (range 0-22.0%). Cystine and struvite (magnesium ammonium phosphate) stones were rare, with 1.0% and 2.2% respectively within the whole cohort.

### Comparison of stone composition and ethnicity

Using a Chi Squared analysis there was no significant difference between stone composition in different ethnic groups (p=0.07) (fig. 1). Figure 1 displays the percentages of each stone type compared to ethnicity. Although the results are not statistically significant, uric acid stones do appear to be more common in the White-other group at 22.0% compared to 10.3% for White British, 5.2% for Asian, 9.52% for Black and 0% for other/ unknown. Calcium oxalate also appeared more common in the Asian population with 71.9% compared with 56.7% in the White British population and 52.6% in the White-other population. Calcium oxalate stones were also common in the Black population at 76.1% and 76.9% in the Other/unknown group however the total numbers for this group were much smaller at 21 and 39 respectively.

**Figure 1.**
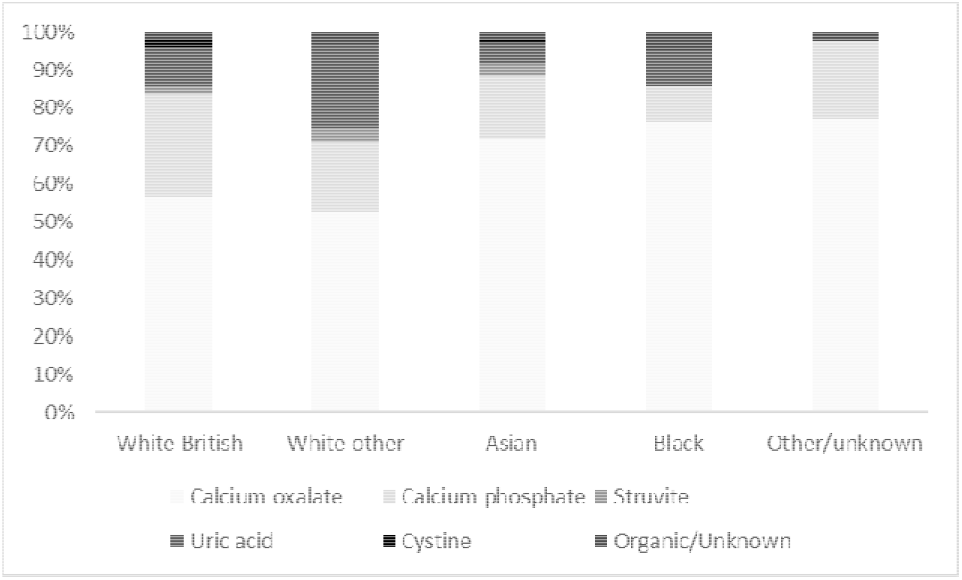
Proportions of different stone compositions compared with ethnicity

### Repeat stone analysis

10% of patients had had a previous stone analysed on our records. A statistically significant number of patients, 87%, formed the same stone before (p=<0.0001 binomial test). Figure 2 shows the results for each stone type. 100% of patients with a calcium oxalate, uric acid and cystine stone had previously formed the same stone. Out of 6 stone formers with calcium phosphate stones, 3 had previously formed calcium phosphate stones, 1 calcium oxalate and 2 struvite. Out of 2 stone formers with struvite stones, 1 formed the same stone and 1 formed a calcium phosphate stone previously.

**Figure 2.**
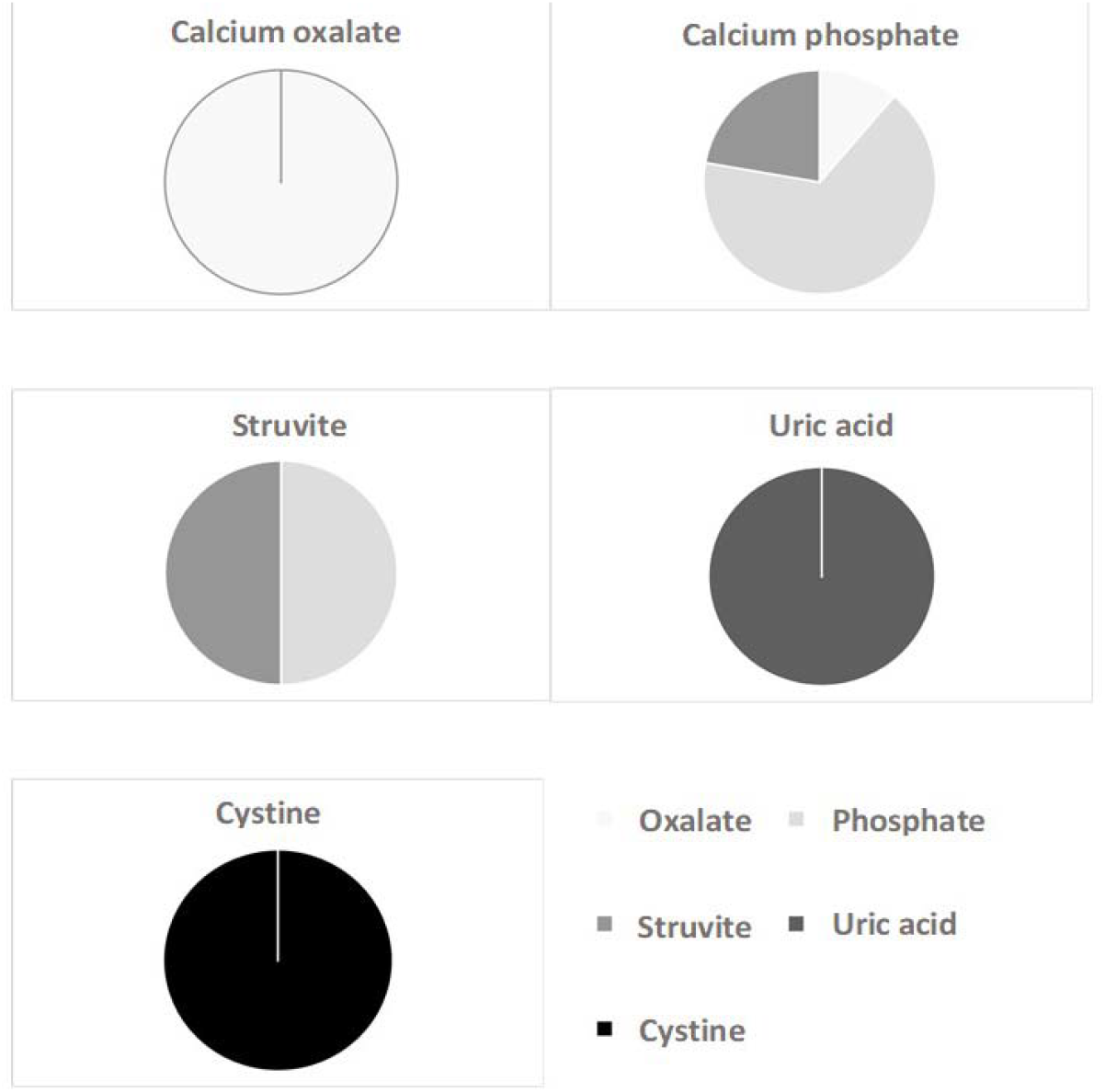
Pie charts for each stone composition: displaying the stone composition for the patients’ previous stones

### Odds ratios of having a kidney stone analysed vs not having a kidney stone analysed for each ethic group

The local population is 48.0% White, 32.5% Asian, 10.0% Black and 9.5% Mixed/ Other[12]. Figure 3 presents the odd ratio for having stone analysis vs not having stone analysis for each ethnic group. The odds for the Black population having a stone analysed (OR 0.62, CI 0.39-0.97, p=0.04) was significantly lower than the local population i.e. the black population in the local area is less likely to get kidney stones than any other ethnic group and is therefore underrepresented in our kidney stone population. The odds for the Asian population having a stone analysed (OR 1.31, CI1.05-1.62, p=0.01) was significantly higher than the local population. There were no statistically significant differences for White (OR 1.09, CI 0.87-1.35, p=0.47) and Mixed/ Other (OR 01.33, CI 0.95-1.87, p=0.09) ethnicities the odds of having a stone analysed compared to local population.

**Figure 3.**
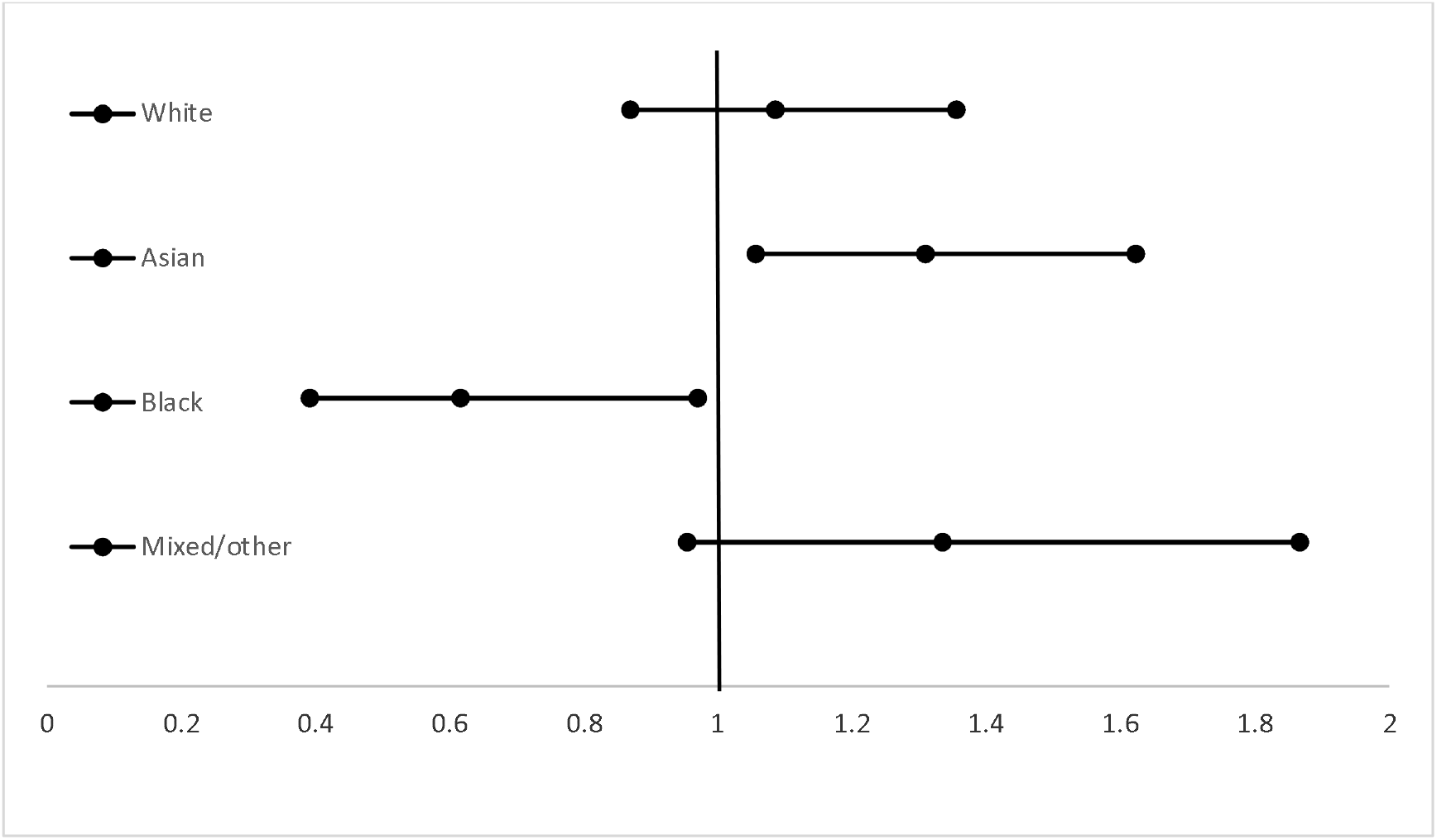
Forest plot for odds ratio of kidney stones by ethnic group compared to local population with 95% confidence intervals

## Discussion

Understanding the epidemiology of stone disease is important for improving disease management. Firstly, it helps us provide an understanding of underlying metabolic disturbances that lead to stone formation. Secondly, we may also be able to predict who will respond to shock wave lithotripsy (SWL) better by knowing the likely composition of the stone. This could reduce the need and therefore cost and radiation exposure of more involved diagnostic tests such as Dual Energy Computed Tomography. Finally, it is vital to guide public health measures which target high risk groups with preventative advice for stone development and recurrence.

### Calcium oxalate stones

The distribution of stone composition observed is similar to the patterns revealed by other studies in the Western world. Two of the largest and most recent studies of stone composition in the Western world are: Daudon et al. 1995[13] and Lieske et al 2014[6]. Daudon analysed 10617 stones, Lieske 43,545 stones: these studies demonstrated a prevalence of calcium oxalate stones of 65.98-67%, compared to 64% in our study.

Our Asian (Indian, Pakistani, Bangladeshi, Chinese or any other Asian background) population had a prevalence of 72% calcium oxalate stones. Most studies in Asia publish slightly higher rates of calcium oxalate stones of 75-93%[1–5]. This may show that our Asian population retain some increased risk of calcium oxalate stones which is slightly moderated by factors unique to a more western location.

### Uric acid stones

In our population the overall rate of uric acid stones was 10% compared to 8-10.8%[6, 7] in other similar studies. However, in our study there appears to be an increased percentage of uric acid stones in the White-other population with 22% uric acid stones. There could be several genetic and environmental factors for this increased risk. Uric acid are associated with a diet rich in meat protein, obesity, metabolic syndrome and insulin resistance[14], any of these could be the cause.

### Recurrent stones

Our patients had a 10% chance of having had a kidney stone analysed within the last 10 years. This is usually quoted as much high due to a meta-analysis of large retrospective studies showing the recurrence rate of renal stones to be 52% at 10 years.[15] We only measured stone which were sent to analysis in one NHS trust. Many stone formers will have had a stone which passed spontaneously and will not have been sent for analysis or will have been treated in a hospital in a different NHS trust or different country. In addition, some patients will have stones operated on where basketing of fragments cannot be achieved or had ESWL on recurrent stones and therefore will not have had a stone sent for analysis. This likely accounts for the lower recurrence rate in our study.

Patients with calcium oxalate, cystine and uric acid stones formed the same stone again 100% of the time. These patients probably each a have a dominant causative factor influencing their stone production. However, patients who had previously formed calcium phosphate and struvite stones could form either of these stones in our study, or a calcium oxalate stone.

### Odds ratios for different ethnicities having stone analysis

As would be expected, we found a lower odds ratio for stones in the black population. These results are concordant with studies done in the United States[9], [17].

Our Asian population was mostly from South Asia with 39.6% Bangladeshi 39.6%, 19.8% Pakistani, 16.7% Indian and 3.1% Chinese and 20.8% from any other Asian background. We would expect a higher stone prevalence in this population as there is a well-known stone forming belt stretching across the West Asia, Southeast Asia, South Asia[18].

## Limitations

A completely accurate picture of the incidence of different stone types in different ethnic populations cannot be derived from this study. The study was not prospective; 39 of our patients did not state their ethnicity or stated their ethnicity as other. In addition, most of our stones have come from patients undergoing percutaneous nephrolithotomy or ureteroscopic removal of stones, with a small minority of stone which were spontaneously passed. Therefore, it does not account for the majority of spontaneously passed or asymptomatic stones.

The patients in this area should have broadly similar environmental factors such as socio-economic status and climate. However, we cannot control for a multitude of confounding factors such as diet, obesity, medications, family history, and medical conditions such as hypertension, gout, inflammatory bowel disease, bone disease and diabetes. We also do not know how long our patients have been resident in this area. We know for example that the population of White-other patients has increased from 8.2% to 12.6% in London between 2001 and 2011^11^. As stones take many years to form some patients may have been exposed to risk factors in a different geographical location for many years before presenting at the hospitals in East London.

These limitations withstanding, our results represent the first study into ethnicity and stone composition in the UK and raises several points of interest that warrant additional study.

## Conclusions

This study allowed for a unique comparison between an ethnically diverse population within the same location. Uric acid stones are found more frequently in the White-other population and calcium oxalate stones are found more frequently in the Asian and black population. The odds ratio for having a stone was significantly higher in the Asian population and lower in the Black population. Repeat stone formers with calcium oxalate, calcium phosphate or cystine stones form the same stone again 100% of the time. Whereas struvite and calcium phosphate stones are correlated indicating that there may be common causative factors.

## Data Availability

All data produced in the present study are available upon reasonable request to the authors

## Acknowledgments

Many thanks to Barts Health NHS Trust.

## Compliance with Ethical Standards

### Research involving Human Participants and/or Animals

No changes were made to patient treatment, this was a retrospective study.

### Informed consent

Patients did not formally consent to this study, however patient information was collected and recorded in accordance to the Data Protection Act. Patients were coded using a unique identifier and no patient identifiable information was recorded, information was kept securely.

